# Dynamics of COVID-19 pandemic at constant and time-dependent contact rates

**DOI:** 10.1101/2020.03.13.20035485

**Authors:** Marek Kochańczyk, Frederic Grabowski, Tomasz Lipniacki

## Abstract

We constructed a simple Susceptible–Infected–Infectious–Excluded model of the spread of COVID-19. The model is parametrised only by the average incubation period, *τ*, and two rate parameters: contact rate, *r*_C_, and exclusion rate, *r*_E_. The rates can be manipulated by non-therapeutic interventions and determine the basic reproduction number, *R* = *r*_C_/*r*_E_, and, together with *τ*, the daily multiplication coefficient at the early exponential phase, *β*. Initial *β* determines the reduction of *r*_C_ required to contain epidemic spread. In the long-term, we consider a scenario based on typical social behaviours, in which *r*_C_ first decreases in response to a surge of daily new cases, forcing people to self-isolate, and then slowly increases when people gradually accept higher risk. Consequently, initial abrupt epidemic spread is followed by a plateau and slow regression. This scenario, although economically and socially devastating, will grant time to develop, produce, and distribute a vaccine, or at least limit daily cases to a manageable number.

## INTRODUCTION

On the 11^th^ of March, 2020, the World Health Organization (WHO) declared Coronavirus Disease 2019 (COVID-19) a pandemic. The rapidly spreading disease is caused by a novel coronavirus, SARS-CoV-2, transmitted between people via respiratory droplets. Typical symptoms are fever, cough, and shortness of breath. In severe cases, infection leads to pneumonia and acute respiratory distress syndrome. There is currently no vaccine or specific antiviral treatment. The virus was first reported in Wuhan, Hubei, China, in December 2019. The disease had spread widely over the province of Hubei, but was contained after strict quarantine imposed on January 23, 2020. The peak of daily confirmed cases of about 4000 was reached on February 3, ten days after the quarantine had been imposed; then the number of daily new cases decreased below 100 at the beginning of March. Currently (as of March 20, 2020), despite some efforts, nearly exponential growth with more than 20 000 cumulative cases is observed in Italy (nearly 50 000 cumulative cases), Spain, Germany, and the US. In contrast, South Korea, that reported less than 10 000 cumulative cases, appears to have exited the exponential phase.

To characterize the dynamics of COVID-19 spread and be able to consider possible long-term scenarios, we developed a simple Susceptible–Infected–Infectious–Excluded model. The model has only 3 parameters: average incubation period (known to be approximately 5 days [1, 2]) and two parameters that can be modified by applying non-pharmaceutical protective measures: the infectious–susceptible contact rate, *r*_C_, and the exclusion rate, *r*_E_. Quarantine reduces *r*_C_, while testing increases the rate at which infectious individuals are hospitalised or isolated, *r*_E_. These two rates determine the basic reproduction number, *R* = *r*_C_/*r*_E_. The model is applied to estimate the fold change of *r*_C_ required to contain the epidemic at its early stage. Since early containment seems implausible, we use the model to put forward longer-term scenarios, in which we assume time-dependent *r*_C_ reflecting typical social behaviours rather than the impact of government-mandated restrictions that appear to be ineffective in European countries.

## RESULTS

### Model

We consider a deterministic **S**usceptible–**I**nfected– **I**nfectious–**E**xcluded (SIIE) model shown in figure 1. Our model is similar to a commonly used SIR model extension, SEIR [3, 4]; key differences and their consequences are explained in Discussion. We assume that an individual can be in one of the following states: *susceptible* (S), *infected* (i_1_, …, i_5_), *infectious* (I), or *excluded* (E). *Susceptible* individuals get infected by contact with *infectious* individuals at the rate proportional to the contact rate, *r*_C_. Multiple *infected* states are introduced to account for the average incubation period, *τ*. With *k* = 5 *infected* states such representation is equivalent to the assumption that the incubation period is distributed according to the Erlang distribution with *k* = 5 and the rate parameter *λ* = *k*/*τ* = 1/day. *Infectious* individuals may become *excluded* with the exclusion rate *r*_E_. As a consequence, the serial interval is distributed according to the hypoexponential distribution that is a series of *k* = 5 exponential distributions, each originating from one step of the incubation process, having mean times *τ*/*k* = 1 day, and one exponential distribution, originating from the “exclusion” process, with mean time 1/*r*_E_ = 3 days. The serial interval distribution implicated by the model agrees perfectly with the distribution obtained from epidemic data (see figure 2B in Ref. [5]). *Excluded* individuals comprise the confirmed cases currently handled by the healthcare, those recovered with acquired resistance, and the deceased. *Excluded* individuals cannot infect. We assume that *excluded* individuals maintain resistance to the disease. The dynamics is governed by a system of 8 ordinary differential equations provided as equations (8a–8e) and analysed in Methods.

**Figure 1.**
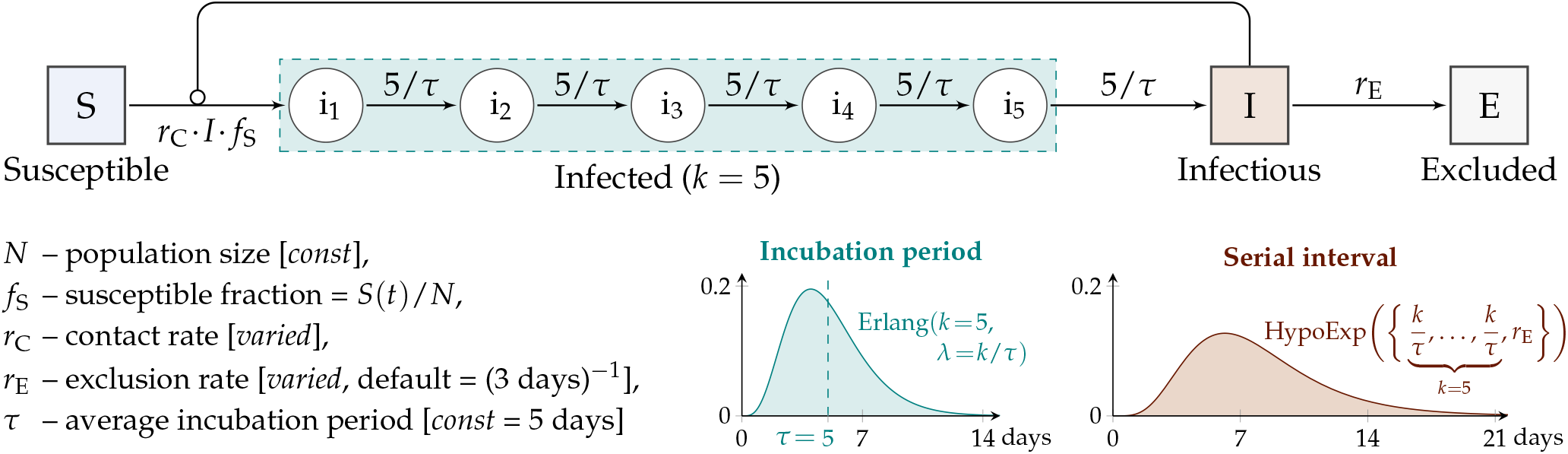
Scheme of the model with notation guide and default parameter values. The incubation period, resulting from inclusion of *k* = 5 intermediate *infected* states, follows the Erlang distribution with average *τ* = 5 days, in agreement with data [1, 2]. The serial interval follows the hypoexponential distribution with rate parameters *λ* = {5/*τ*, 5/*τ*, 5/*τ*, 5/*τ*, 5/*τ, r*_E_}. The model is fully characterized by three parameters: *r*_C_, *r*_E_, and *τ*.

**Figure 2.**
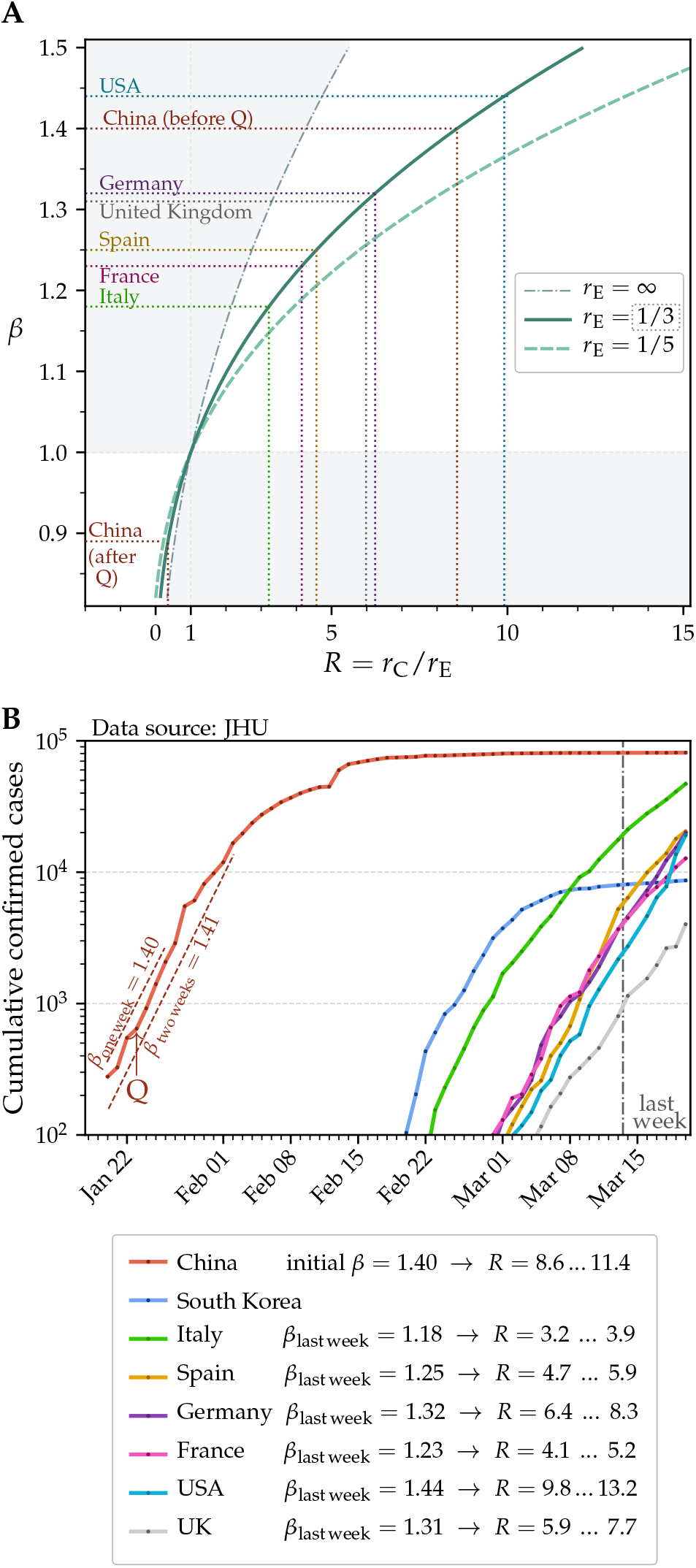
Daily multiplication coefficient predicted by the model and estimated from data. (**A**) Dependence of the daily multiplication coefficient, *β*, on the basic reproduction number *R* = *r*_C_/*r*_E_. Dotted lines guide the eye to relate *R* and *β* coming from data for China before and after quarantine (marked with ‘Q’) and for several European countries and the US based on last-week data. (**B**) Epidemic dynamics in selected countries with high number of cumulative cases of COVID-19 based on data from Johns Hopkins University (JHU) [6]. Value of initial *β* for China is estimated based on 7 or 14 days of early exponential phase; current *β* is estimated for countries being in the exponential phase based on last-week data (7 days since March 14 to March 20, 2020). The range of values of *R* is computed assuming that 1/*r*_E_ is in between 3 and 5 days.

The exponential phase of the epidemic growth, during which the number of *excluded* (confirmed) individuals grows as *E*(*t*) = *E*(*t* = 0) *×* exp(*α t*), can be analysed based on an implicit formula following from equation (9) (see Methods):

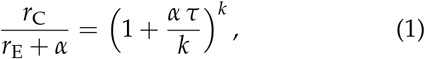

which can be replaced by

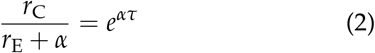

when the number of intermediate steps *k* → ∞.

### Dynamics of infection spread with respect to the contact rate and the exclusion rate

In the subsequent analysis we will use the daily multiplication coefficient, *β*, which is the factor that describes the day-to-day increase of the new confirmed cases (for example, when the number of new cases is 25% higher than the number of new cases on the previous day, then *β* = 1.25). We use equation (1) to calculate the dependence of *β* = exp(*α*) on *r*_C_, *r*_E_, and *τ* (supplementary figure S1). Either a decrease of the contact rate *r*_C_ or increase of the exclusion rate *r*_E_ may result in reduction of *β* below 1 and containment of the epidemic. Practically, *r*_C_ is reduced by quarantine or isolation of individuals, which both lower the potential number of contacts per individual per day, while *r*_E_ is increased by performing more diagnostic tests, which reduces the expected time during which an infected individual may infect others. Coefficient *β* decreases also with increasing *τ* (supplementary figure S1C), but this parameter is not controllable by protective measures.

Equations (1) and (2) imply that

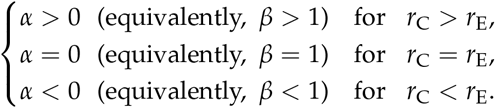

Further, parameters *r*_C_ and *r*_E_ determine the expected number of *susceptible* individuals that will be infected by each *infectious* individual, *R*:

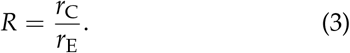

Although the daily multiplication coefficient, *β*, is not a function of solely *R*, the value of coefficient *R*, being equivalent to the commonly used basic reproduction number (usually denoted ℛ_0_), is critical for propagation of the epidemic: *R* = ℛ_0_ *>* 1 implies exponential progression, *R* = ℛ_0_ *<* 1 implies exponential regression.

It is important to notice that the number of *excluded* individuals may be much lower than the total number of *infected* and *infectious* individuals: *i*_1_ + …+ *i*_*k*_ + *I*. The confirmed fraction

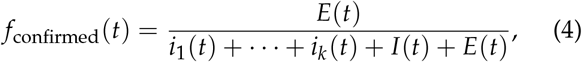

that remains constant in time in the exponential phase, can be calculated based on equations (10) and (11) as

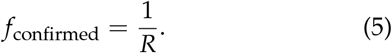

The epidemic growth that is associated with large *R* implies a proportional large number of not yet confirmed cases that in the next few days will contribute to the number of cumulative confirmed cases.

In figure 2A we show *β*(*R*) for two values of *r*_E_: 1/5 and 1/3 day^−1^, and in the limit of *r*_E_ → ∞. The figure should be read as follows. An initial *β* that can be determined based on timeline data for a considered country or region corresponds to some value of *R*. Since to terminate the exponential growth phase, *R* has to be reduced to 1, figure 2A shows how much *r*_C_/*r*_E_ = *R* should be reduced to contain the epidemic. The reproduction number can be reduced by either reducing the contact rate or increasing the exclusion rate, but the latter is problematic when the number of cases grows rapidly.

Assuming *r*_E_ = 1/(3 days), which is the default value in the model, the data showing epidemic dynamics in China (figure 2B) can be interpreted as follows. Based on the initial exponential phase (trends in one or two weeks marked in figure 2B) one can estimate that *β* = 1.4 which corresponds to *R* ≃ 8.6 (figure 2A). Thus, to “stabilise” the epidemic, China had to reduce *R* by a factor of 8.6. However, as indicated by the fall of the number of daily new cases, China reduced *β* to approximately 0.89 = *β*′ (see Methods), which implies *R* = 0.34 and effective reduction of *R* about 25 times. We should notice that several authors [7, 8], also based on epidemiological data from China, obtained much lower estimates for *R*; we will come back to this important issue in Discussion.

The ∼10-day time lag between introduction of the quarantine and the time of the peak number of daily new cases can be attributed to: (1) the incubation period that causes that infected individuals are registered about *τ* = 5 days after infection and (2) the fact that the quarantine does not prevent mixing of infectious and susceptible individuals in households, adding next *τ* = 5 days to the onset of the disease among family members. Unfortunately, this implies that even if strict quarantine is imposed immediately in European countries it will have a delayed effect.

In the case of Italy, soft quarantine imposed on February 21 in the most affected Italian province, Lombardy, allowed to reduce *β* from the initial value of 1.5 (calculated based on the week of February 23–29, 2020) to the current value of *β*_last week_ = 1.18 (calculated based on the last week, March 14–20, 2020). This implies reduction of *R* from about 12 to 3, which is only about 4 times. To just contain the epidemic, that is, reduce *R* to 1 and stabilise the number of daily cases, Italy should *further* reduce the contact rate three-fold. Figure 2A indicates that four other large European countries (Germany, France, Spain, and United Kingdom) as well as the US, each having thousands of cases and *β*_last week_ *>* 1.2 (as of March 20, 2020), to contain the epidemic should reduce the contact rate at least four-fold. In figure 2B we give current values of *β* and corresponding values of *R* for the mentioned countries [estimated for *r*_E_ = 1/3 (default value in the model) and *r*_E_ = 1/5 day^−1^].

A much different dynamics can be observed in South Korea, which is exiting from the exponential phase (figure 2B). Among many preventive countermeasures, until March 20, 2020, South Korea performed 316 000 tests [9], likely substantially increasing the exclusion rate. The example of South Korea is promising as it suggests that development of easily accessible, fast, and accurate tests may help to contain the spread of the disease.

### Hypothetical long-term scenarios

The above analysis of the epidemiological situation in several European countries and in the US suggests that the administrative protective measures adopted in these countries failed to contain the epidemic. It is thus worth to consider long-term scenarios in which the cumulative number of *excluded* cases, *E*, becomes comparable to the population size, *N*, substantially decreasing the susceptible fraction, *f*_S_, over time. In such case, initial exponential growth ceases and the number of daily new cases reaches a maximum.

When the reproduction number remains constant, the maximum number of new daily cases is fully determined by initial *β* and exceeds 0.3 million daily new cases for *β* = 1.05 and 1 million cases for *β* = 1.1 in a population of *N* = 50 million people (supplementary figure S2A). It is clear that even this smaller number exceeds the health-care capacity of a country of tens of millions inhabitants such as Italy (∼ 60 million), South Korea (∼ 50 million), or Poland (∼40 million). This, in turn, implies higher case fatality rate, which may approach the rate of severe cases (10–15% [10]). Correspondingly, with *β* changing from 1.05 to 1.1, time to reach the maximum of daily new cases shortens nearly twice: from 7.5 to 4 months, assuming the initial number of *excluded* (confirmed) cases, *E*(*t* = 0), equal 500. We should notice that *β* = 1.05 as well as *β* = 1.1 require quarantine; in “non-quarantined” countries, such as currently France, Germany, Spain, United Kingdom, and the US, *β >* 1.2 (figure 2B).

In supplementary figure S2B, we simultaneously show the number of the maximum weekly and monthly new cases and the time to reach the maximum. Parameter *β >* 1.1 implies that within one (worst) month more than half of population will be infected, whereas *β <* 1.1 require long-term quarantine.

We consider the scenario with the constant reproduction parameter rather not realistic. Initially, when the number of cases is relatively low, the only way to reduce the contact rate is an administratively-imposed quarantine. However, one could expect that when the number of daily new cases exceeds a threshold, people will begin reducing their contacts voluntarily [11] or taking other protective measures such as careful hand-washing or physical distancing to reduce their number of effective contacts (reviewed in Ref. [12]). To account for this behavioural aspect, we assume that the contact rate is a decreasing function of daily new cases. We propose to assume that

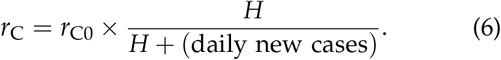

This simple formula implies, in particular, that when the number of daily new cases equals *H*, people reduce their contacts twice. The value of parameter *H* can be culture-dependent. In figure 3A we show long-term epidemic dynamics assuming *H* = 10 000. In the considered scenario, the number of daily new cases relatively quickly approaches 15 000 and then decreases slowly over years with a decreasing susceptible fraction, *f*_S_. The scenario depicted in figure 3A results in a manageable number of new cases and grants time to develop, produce, and distribute a vaccine.

**Figure 3.**
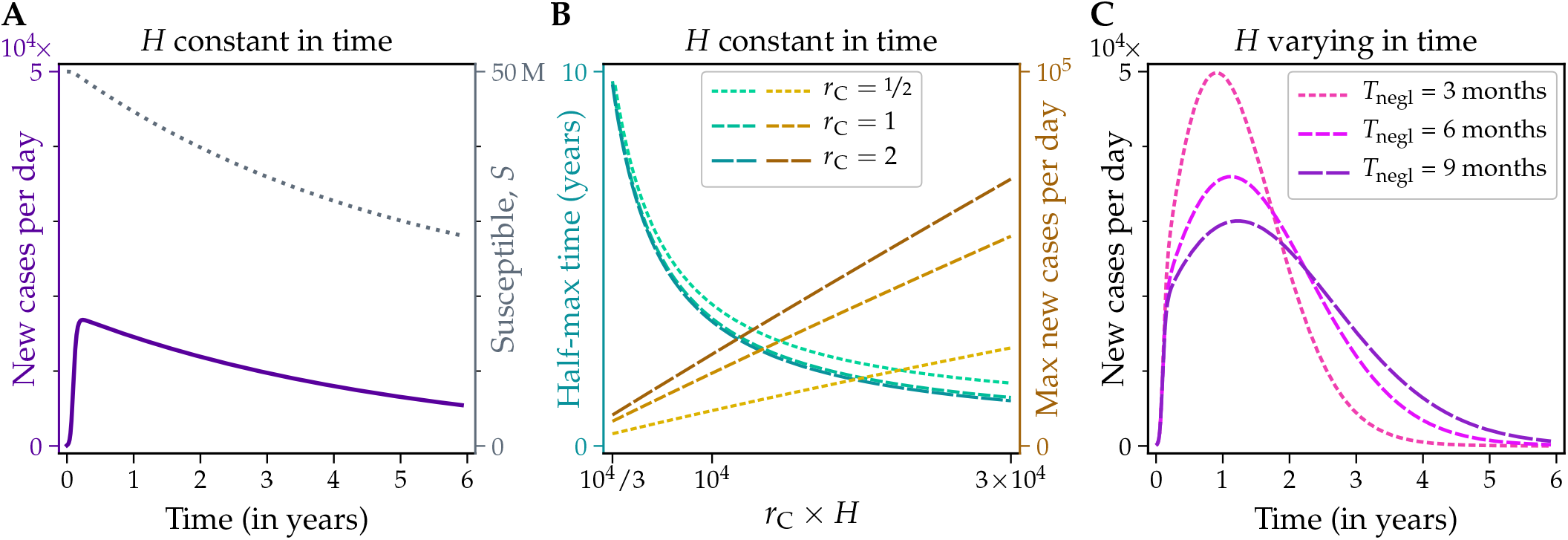
Long-term epidemic dynamics with time-dependent contact rate. (**A, B**) Contact rate reduced in response to the surge in the daily number of cases. (**C**) Contact rate reduced in response to the surge in the daily number of cases and modulated by the duration of the quarantine. In panels A and C, initial *β* = 1.18, which is equal to *β*_last week_ for Italy. In all panels, population size *N* = 50 million and *E*(*t* = 0) = 500.

In this scenario, for a given *r*_C0_, the maximum number of daily cases is an increasing function of *H*, whereas the time in which the number of daily new cases decreases to the half of the maximum value, *t*_↘max/2_, decreases with *H*. A bit surprisingly, *t*_↘max/2_ depends mainly on *r*_C0_ *× H*.

Finally, we consider a scenario in which a secondary behavioural effect modifies the primary effect discussed above. As before, we assume that people reduce their contacts in response to an increasing risk, but in the course of their (self)quarantine they gradually accept an increasing risk [13]. (As an aside, when protective measures are lifted rapidly, such behaviour can lead to epidemic waves observed during the 1918–1919 H1N1 influenza pandemic [14].) Here we propose to model this effect by assuming that

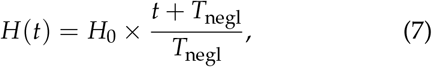

where *t* is (somewhat arbitrary) time from the beginning of the epidemic and *T*_negl_ is the time scale on which *H* (and risk acceptance, or “negligence”) doubles. In figure 3C we show long-term epidemic dynamics assuming *H* = 10 000, and three values of *T*_negl_: 3, 6, and 9 months. Interestingly, the dynamics is characterized by a relatively broad plateau, which is desirable due to limited capacity of the healthcare system.

## DISCUSSION

In this study, we proposed a Susceptible–Infected– Infectious–Excluded model to analyse how the dynamics of the COVID-19 spread depends on the contact rate, *r*_C_, and the exclusion rate, *r*_E_—two model parameters that can be modulated by non-therapeutic interventions. The remaining model parameter is the average incubation period, assumed equal 5 days [1, 2]. A decrease of the contact rate, that can be achieved by imposing quarantine, results in a decrease of the daily multiplication coefficient *β* that describes growth of the cumulative as well as the daily new cases in the exponential phase. Coefficient *β* can be reduced also by an increase of the exclusion rate that in turn depends on efficiency of the healthcare. Large exclusion rate is achieved by fast identification and isolation of *infectious* individuals. However, when the number of cases is very high, achieving large exclusion rate is nearly impossible; in this case, reduction of the contact rate remains the only option.

In the early phase, before preventive measures were introduced, COVID-19 had been spreading in China and then in Italy with *β* = 1.4 and *β* = 1.5, respectively. This allowed us to estimate the initial basic reproduction number in these countries: *R* ≃ 9 and *R* ≃ 12, respectively. Of note, even without prevention the reproduction number depends on numerous different factors such as sociological context, hygiene, and weather.

In several important studies (conveniently summarized on a webpage of an online SEIR model simulator [15]), the estimated initial reproduction number in China was in the range 2–7, which is lower than in our estimates. In particular, Kucharski *et al*. [7] found initial *R* = 2.35 (95% CI: 1.15–4.77), while Tang *et al*. [8] found *R* = 6.47 (95% CI: 5.71–7.23). Both estimates were based on the SEIR model, which is essentially very similar to our SIIE model. In SEIR model, **S**usceptible individuals become **E**xposed, then **I**nfected (which correspond to our *infectious*), and then **R**emoved (which correspond to our *excluded*). The main difference between our model and the SEIR model lies in the fact that instead of a single *infected* state we have assumed the existence of five intermediate *infected* states, which is equivalent to assuming that the incubation period is Erlang-distributed with parameters *k* = 5 and *λ* = *k*/*τ*, while serial interval follows the hypoexponential distribution, in agreement with Li *et al*. [5]. Importantly, the simplifying assumption that there is only one intermediate state is equivalent to the assumption that the incubation period follows an exponential distribution. A bit surprisingly, this simplification substantially modifies *R* estimated from *β*. In figure 4, we show *β*(*R*) for two values of *r*_E_: 1/3 (default) and 1/5 day^−1^, calculated assuming that the number of intermediate states is 1, 2, 3, 5, 10 or ∞ (the last case has been determined based on an equivalent model represented by delay differential equations, see Methods). One can observe that for *β* = 1.4, the value of *R* predicted for a model with one intermediate state is about twice smaller than *R* predicted for the model with *k* = 5 intermediate states. The exponential distribution or even Erlang(*k* = 2, *λ* = *k*/*τ*) distribution of the incubation period (figure 4C) is not supported by data [1, 2], which indicates much narrower distribution isolated from zero, so we think that our estimate of the basic reproduction number is closer to a real value.

**Figure 4.**
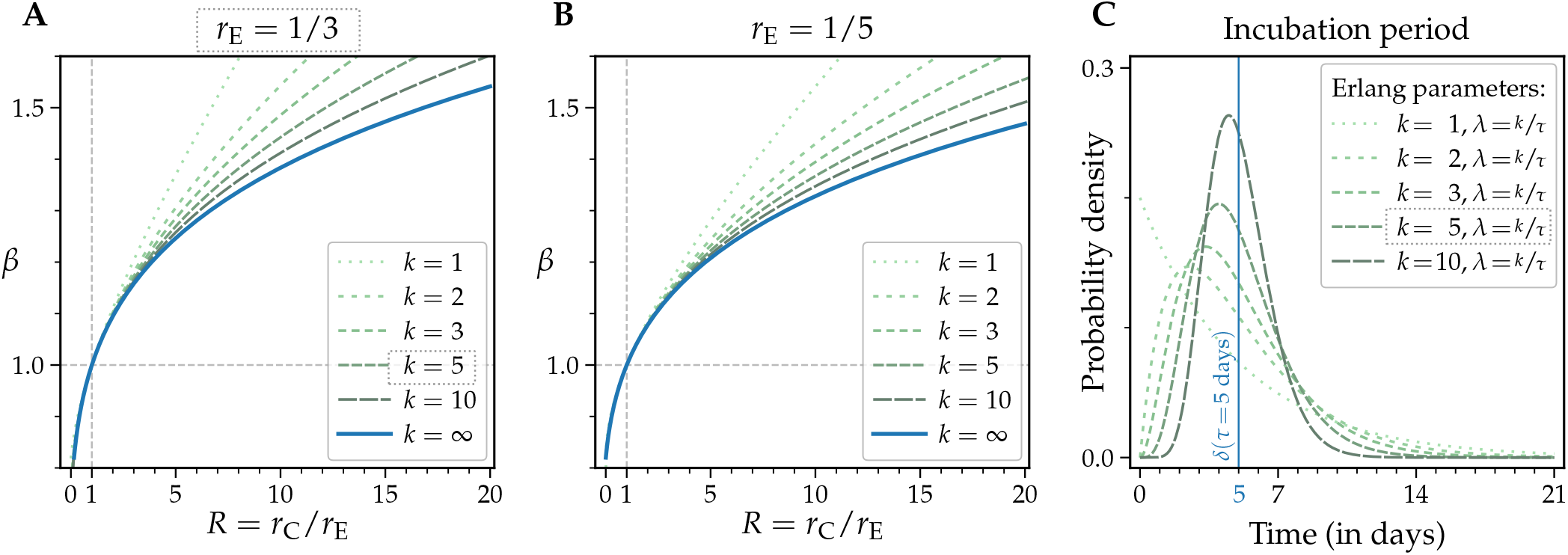
Dependence of (A, B) the daily multiplication coefficient, *β*(*R*), and (C) the distribution of the incubation period, *τ*, on the number of *infected* intermediates.

Data describing epidemic dynamics in China suggest that very strict quarantine and other preventive efforts imposed in the most affected China province, Hubei, [7, 16] allowed to reduce *R* about 25-fold (which is on par with the estimated reduction of transmissibility of 97– 100% [17]). This allowed the Chinese to reduce coefficient *β* from about 1.4 in the exponential growth phase to about *β*′ = 0.89 in the regression phase. In Italy, however, in response to the mild restrictions, *β* was reduced from about 1.5 in the early phase to 1.18 observed currently (March 14–20, 2020), which corresponds to the reduction of *R* from about 12 to 3; thus, further three-fold reduction is necessary to contain the exponential epidemic spread.

Higher reduction of *R*, at least four-fold, is required in four other considered European countries, France, Germany, Spain, and United Kingdom, with the number of cases exceeding 10 000, and current *β >* 1.2 (March 14– 20, 2020). Such reduction of *R* would allow only to limit growth of daily new cases, but further four-fold reduction is necessary to bring the epidemic to the regression phase with *β*′ = 0.89, as was done in Hubei. Higher *β*′ ∈ (0.89, 1.0) may also allow to eliminate the disease but the required quarantine time would be longer. It appears that most of European countries follow the regulations adopted in Italy, which proved to be insufficient to contain the epidemic, and, correspondingly, with a week-to-month delay, follow the Italian trajectory of the cumulative daily new cases.

Of concern is COVID-19 lethality, which in quantitative terms is usually expressed as the case fatality rate, assessed as the ratio of deaths to confirmed cases. This definition, used by WHO, works well in the steady state when the number of active cases remains constant or when historical data of previous epidemics are analysed. However, in the exponential growth phase, the case fatality rate underestimates the real rate of deaths due to the disease. When the average time between the onset of symptoms or positive test and death is *m* days, the current cumulative number of deaths should be divided by the cumulative number of cases recorded *m* days before, which is *β*^*m*^ times lower than the current number. This observation may to some extent explain puzzling large differences between fatality rates across European countries. In Germany, having the lowest fatality rate (among countries with a high number of cases), identification of people by testing at early stage of the disease and high standard of public healthcare may result in large *m*, which together with high *β* ≈ 1.3 (March 14–20, 2020) may result in a significant underestimation of the case fatality rate. Assuming *m* between 10 and 14 days, one can find that the case fatality rate may be underestimated by a factor between 1.3^10^ and 1.3^14^, that is, 14–40 times. Finally, we should notice that in non-containment scenarios the expected death toll may not be predicted based on the case fatality rate, because a significant number of cases may be asymptomatic (and some individuals could possibly turn out resistant).

Since current data suggest that in Europe, US, and Iran the epidemic will not be contained in its early phase, we have analysed longer-term scenarios. The model with the constant contact and exclusion rates predicts that even for a relatively small exponential growth phase coefficient *β* = 1.05, the number of daily new cases (in a 50-million population) will exceed 0.3 million, well above the capacity of the healthcare system. Additionally, keeping the coefficient *β* at such low level requires stringent limitation of personal contacts, while the time to the peak of the epidemic would be about 7 months. One of potential consequences of exceeding the healthcare system capacity is the increase of case fatality rate due to the lack of necessary medical equipment.

We think that the more realistic scenarios are these in which the contact rate varies over time. In early phase of the epidemic, the contact rate may be reduced only by forcing people to stay at home; in the latter phase, when the number of daily cases exceeds a threshold, people isolate themselves to reduce the risk. In such scenario the number of daily new cases reaches peak proportional to an assumed “fear” threshold and then slowly decreases due to the decreasing fraction of susceptible individuals. Such scenario seems more realistic and, although devastating for both the economy and social life, grants time to develop and administer vaccine. Historical data on 1918–1919 H1N1 influenza pandemic suggest also that this “fear” threshold may not be constant in time, because people suffering from prolonged quarantine may tend to accept higher risk. When this “negligence” effect is included, one may obtain trajectories for which fast growth is followed by a plateau and then relatively fast decrease of daily cases. A bit surprisingly, this scenario, although not resulting from centrally imposed preventive policies, may be the most plausible non-containment scenario balancing health and economic costs.

## METHODS

### Model equations

The dynamics of the system depicted in figure 1 is governed by the following system of 8 ordinary differential equations (ODEs):

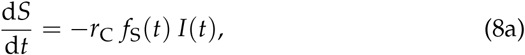

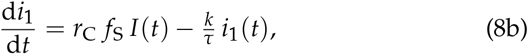

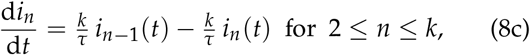

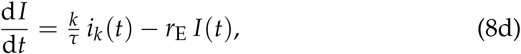

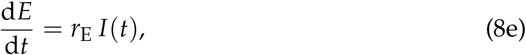

where *k* = 5 is the assumed number of intermediate infected states and *f*_S_ = *S*(*t*)/*N* with *N* = *i*_1_(*t*) + … + *i*_*k*_(*t*) + *I*(*t*) + *E*(*t*) = const being the population size (with included deaths).

In the early phase of the epidemic, *f*_S_ ∼ 1. Under the assumption that *f*_S_ = 1, the systems has the exponential solution:

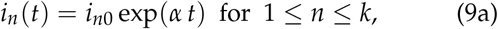

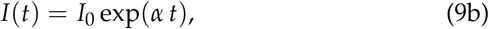

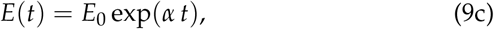

with *E*_0_ being the initial cumulative number of known cases and

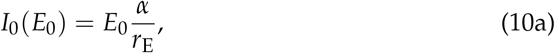

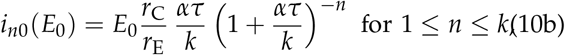

where *α* is given implicitly:

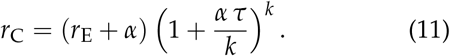

When *N* − *S*(*t*) ≪ *N* is not satisfied, the system becomes essentially nonlinear and has no analytical solutions. This system is solved numerically using Wolfram Mathematica (Wolfram Research, Inc., Champaign, IL, US; notebook is available online [18]). All numerical simulations start from an initial condition in which *i*_*n*_(*t* = 0) = *i*_*n*0_, *I*(*t* = 0) = *I*_0_, *E*(*t* = 0) = *E*_0_, and 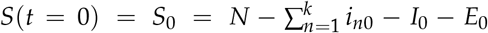, where *E*_0_ is the given initial cumulative number of known cases and *i*_*n*0_(*E*_0_) and *I*_0_(*E*_0_) are computed according to equations 10, assuring that the initial condition lies on the exponential trajectory. In the model variant with varied contact rate, *r*_C_ = *r*_C0_ *× H*/ (*H* + (daily new cases)), daily new cases are computed as *r*_E_ *I*(*t*). Simulations of this model variant start from the same initial condition that lies on the exponential trajectory.

It is noteworthy that in the limit of *k* → ∞ and in the phase of epidemic progression, the considered model is equivalent to the following three-compartment model with delay *τ*:

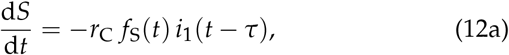

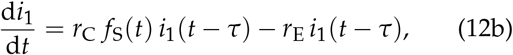

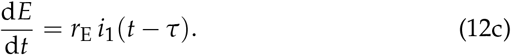

with *f*_S_(*t*) = *S*(*t*)/*N* where *N* = *S*(*t*) + *i*_1_(*t*) + *E*(*t*) = const. One should notice that delay differential equations (DDEs)-based formulation [equations (12a–12c)] may admit negative-valued solutions in the epidemic regression phase.

### Estimation of *β*

Parameter *β* for the initial exponential growth case is estimated based on JHU data [6] of cumulative cases as:

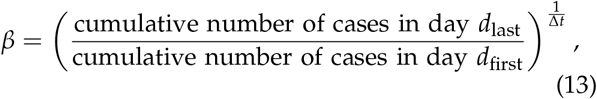

where for China day *d*_first_ is January 20 and day *d*_last_ is January 26 or February 2, 2020, with Δ*t* = *d*_last_ − *d*_first_ – 1 = 6 or 13, respectively. For other countries, last-week data are used: *d*_first_ is March 14 and *d*_last_ is March 20, 2020; Δ*t* = 6.

It should be noticed that as long as the epidemic progresses exponentially, coefficient *β* estimated based on the increase of cumulative cases *E*(*t*) and coefficient *β*′ estimated from the number of daily new cases, *E*(*t*) − *E*(*t* − 1 day), should be equal (which can be obscured by unavoidable fluctuations in daily data). However, when the epidemic ceases, *β* calculated based on *E*(*t*) reaches 1, while *β*′ becomes smaller than 1, reflecting an exponential fall of the number of the daily new (as well as the active) cases. Parameter *β*′ in the epidemic regression phase in China has been estimated as the decay rate of the daily new cases in the time span from February 2 to March 2, 2020.

## Data Availability

Data referred in the manuscript are available at
JHU CSSE, https://systems.jhu.edu/research/public-health/ncov, accessed: 2020-03-20
and
https://ourworldindata.org/covid-testing, accessed: 2020-03-20

http://pmbm.ippt.pan.pl/model/covid19

## SUPPLEMENTARY DATA

**Figure S1.**
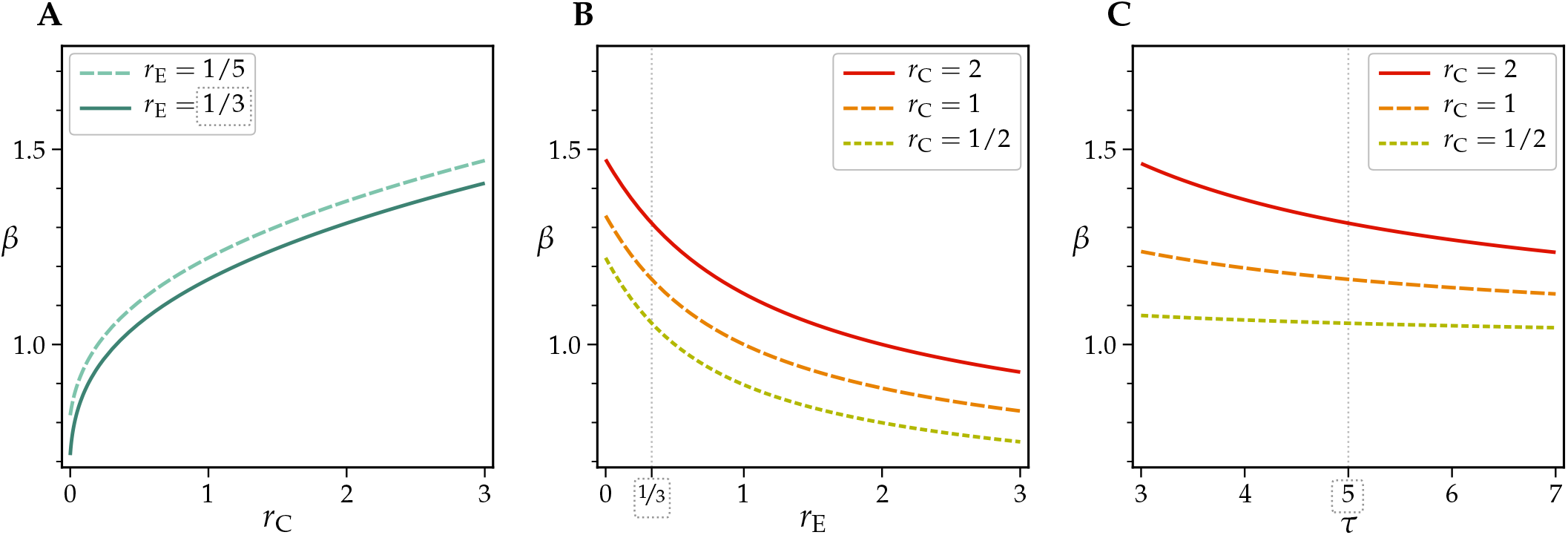
Dependence of the daily multiplication coefficient, *β*, on all model parameters: (A) contact rate, *r*_C_, (B) exclusion rate, *r*_E_, and (C) average incubation period, *τ*. Default parameter vales are marked with dotted rectangles.

**Figure S2.**
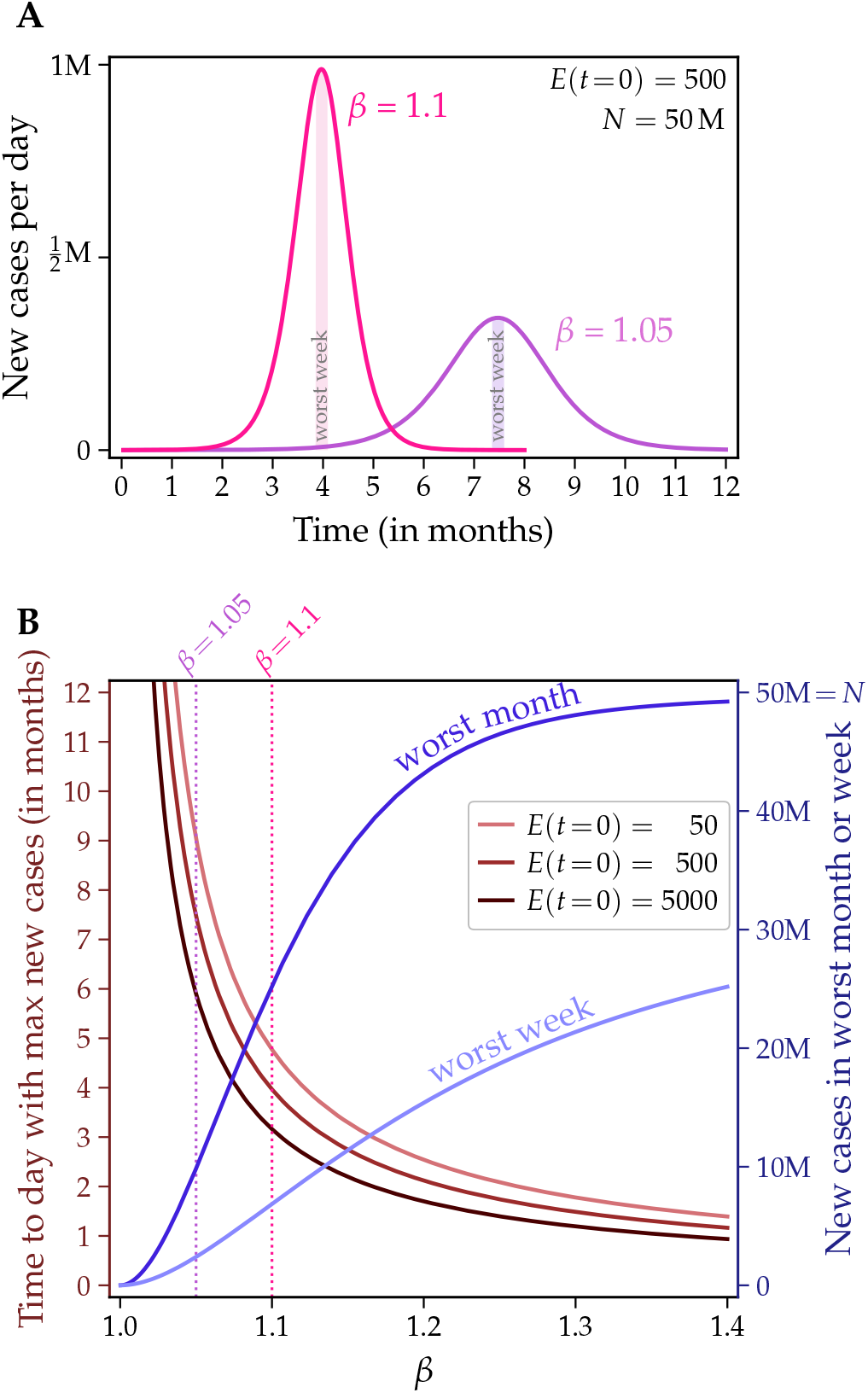
Long-term epidemic dynamics at a constant contact rate. (**A**) Daily new cases for two values of the initial daily multiplication coefficient, *β*, and the initial number of *excluded* (confirmed) cases, *E*(*t* = 0), equal 500. (**B**) Time to the worst day for three initial numbers of confirmed cases and new cases in the worst week and month. In both panels, population size *N* = 50 million, 1/*r*_E_ = 3 days.

## Notes

### Competing Interest Statement

The authors have declared no competing interest.

### Funding Statement

This study was supported by the National Science Centre (Poland) grant 2018/29/B/NZ2/00668

